# Food-Group Intakes and Depression in Tangerang, Indonesia: An Exploratory Case–Control Study

**DOI:** 10.64898/2026.11.05.26367333

**Authors:** Darien Alfa Cipta, Alvin Saputra, Neneng Suryadinata, Rohana Uly Pradita Siregar, Dwi Savitri Rivami

**Affiliations:** Faculty of Medicine, Universitas Pelita Harapan

**Keywords:** Depression, Food Groups, Sweetened Beverages, Body Mass Index, Case–Control Studies, Indonesia

## Abstract

**Background:** Diet may be associated with depression, but evidence on locally consumed food groups in Indonesia is limited. Objectives: To compare food-group intake between people with and without a formal diagnosis of depression in Tangerang, Indonesia, and to explore whether BMI modifies any association.

**Methods:** In this case–control study, cases were psychiatric outpatients with a formal diagnosis of depression. Controls had no formal diagnosis and were recruited through university and community networks. Daily intake of ten food groups was estimated with a semi-quantitative food frequency questionnaire. Intakes were compared with independent-samples t-tests (Welch’s test when variances were unequal), overall and stratified by BMI group. Logistic regression estimated the odds of depression per 100 g/day of sweetened beverage intake, adjusted for BMI group, sex, and age, with a sweetened beverage × BMI group interaction term.

**Results:** There were 257 participants: 104 cases and 153 controls. Sweetened beverages were the only food group whose intake differed significantly between groups (approximately 214 vs. 153 g/day; mean difference 61 g/day; 95% CI 18–103; p=0.005). Each additional 100 g/day was associated with higher odds of depression after adjustment (OR 1.29; 95% CI 1.09– 1.52). The association appeared stronger in underweight/normal-weight participants (ratio of ORs for overweight/obese vs. underweight/normal 0.70; 95% CI 0.49–0.99; p for interaction=0.046).

**Conclusions:** Higher sweetened beverage intake was associated with depression in this sample, and there was modest evidence that the association differed by BMI group. Because the study is cross-sectional in timing, the case–control samples were selected differently, and many comparisons were made, these findings are exploratory. They do not establish causality.

## INTRODUCTION

Diet is an important area of investigation in depression because it encompasses potentially modifiable behaviors and reflects social and economic circumstances that influence health. A systematic review and meta-analysis of observational studies found inverse associations between several healthy dietary indices and depressive outcomes, with the strongest longitudinal evidence involving Mediterranean dietary adherence^1^. These findings support further investigation of dietary patterns and their components but do not establish that individual foods prevent or cause depression.

Food-group analysis provides a practical way to characterize dietary intake while retaining distinctions between commonly consumed foods and beverages. Cereals and tubers, legumes and nuts, animal-source foods, fruits, vegetables, dairy products, prepared foods, and beverages represent different dietary domains. Their associations with depression may differ, and a finding for one group should not automatically be attributed to a particular nutrient, processing method, or overall dietary pattern.

Potential pathways linking diet and mental health include inflammation, oxidative stress, metabolic regulation, and microbiota–gut–brain signaling. However, much mechanistic evidence remains preclinical, and the proposed pathways interact rather than operate through a single dietary component^2^. Depression and its treatment may also influence appetite, food preferences, and eating behavior. Dietary intake measured after diagnosis may therefore be an antecedent, consequence, or correlate of illness.

Understanding these relationships requires locally relevant evidence. In Tangerang, Indonesia, dietary assessment among psychiatric outpatients and comparison participants recruited through university and community networks provides an opportunity to examine food-group differences associated with depression. The present study analyzed one participant sample rather than treating individual dietary exposures as separate studies.

The objectives were to compare the daily intake of ten food groups between participants with and without a formal diagnosis of depression and to explore the association between sweetened beverage intake and depression after adjustment for BMI group, sex, and age. Additional exploratory analyses examined whether this association differed between underweight or normal-weight participants and those who were overweight or obese.

## METHODS

### Study design and setting

This case–control study was conducted in Tangerang, Indonesia, encompassing Kota Tangerang, Kabupaten Tangerang, and Kota Tangerang Selatan. Data were collected in 2025. Cases were recruited from psychiatric outpatient services at Siloam Hospitals Lippo Village and Siloam Hospitals Kelapa Dua. Controls were recruited through Universitas Pelita Harapan and community networks.

### Participants and depression status

The analytic sample included 257 participants: 104 cases and 153 controls. Eligible participants were aged 18-59 years, were able to understand and complete the questionnaires, and gave informed consent. Cases were defined as outpatients with a formal, primary diagnosis of a depressive disorder according to ICD-10 F32/F33 criteria. Diagnoses were established clinically by board-certified psychiatrists at the participating hospitals. Participants with primary psychotic disorders, bipolar disorders, or active substance use disorders that could independently drive severe dietary alterations were excluded. Controls were defined as participants without a formal psychiatric diagnosis, recruited from the community and university networks.

### PHQ-9 assessment

Depressive symptoms were assessed using the Patient Health Questionnaire-9 (PHQ-9), which covers symptoms during the preceding two weeks.^3^ Formal diagnosis was used to define case–control status. A low symptom score was not interpreted as sufficient evidence to invalidate a recorded diagnosis, and a higher score was not treated as equivalent to a formal diagnosis.

### Dietary assessment and food-group definitions

Dietary intake was assessed using semi-quantitative food frequency questionnaires covering the preceding month.^5^ Reported consumption frequencies and food-specific portion sizes were converted into estimated average daily intake. Intake was expressed in units of 100 g/day. Thus, an intake value of 2.0 corresponded to approximately 200 g/day. Total daily energy intake (kcal/day) was estimated from the FFQ using the *Tabel Komposisi Pangan Indonesia* (TKPI).^6^

Ten food groups were examined:

1. Cereals and tubers.
2. Legumes and nuts.
3. Meat, poultry, and eggs.
4. Fish and fish products.
5. Vegetables.
6. Fruits.
7. Milk and dairy products.
8. Oils and fats.
9. Prepared foods, cakes, and snacks.
10. Sweetened beverages.

### Other variables

Participant characteristics included age, sex, education, height, weight, and BMI. Education was summarized as high school completion or at least a bachelor’s degree. Height and weight were measured by trained enumerators using calibrated equipment. BMI was calculated as weight in kilograms divided by height in meters squared. BMI categories followed the WHO Asia-Pacific classification: underweight <18.5, normal 18.5–22.9, overweight 23.0–24.9, and obese ≥25.0 kg/m^2^. For stratified analyses and regression models, these were combined into two groups: underweight/normal weight and overweight/obese.

### Statistical analysis

Categorical variables were summarized as counts and percentages, and continuous variables as means and standard deviations. Food-group intakes were compared between cases and controls using independent-samples t-tests. Welch’s test was used when Levene’s test indicated unequal variances at p<0.05. Mean differences were calculated as case minus control, with 95% CIs.

Food-group comparisons were repeated separately within the underweight/normal-weight and overweight/obese groups. The distribution of depression status across BMI groups was examined using a chi-square test.

Exploratory logistic regression models examined the association between sweetened beverage intake and depression status, with controls as the outcome reference group. Three models were fitted:

- Model 1 included sweetened beverage intake alone.
- Model 2 included sweetened beverage intake, BMI group, sex, age, educational attainment, and total energy intake (kcal/day). The inclusion of education and total energy intake was specifically designed to adjust for baseline socioeconomic differences arising from the recruitment strategy and to isolate the specific association of the food group from generalized hyperphagia or higher caloric throughput.
- Model 3 added a sweetened beverage × BMI group interaction term to Model 2.

Analyses were performed using IBM SPSS Statistics 30.0. A two-sided p<0.05 was considered nominally statistically significant.

### Ethics

The study was conducted in accordance with the Declaration of Helsinki. Ethical approval was obtained from Faculty of Medicine, Universitas Pelita Harapan. All participants provided written informed consent before data collection. RESULTS AND DISCUSSIONS

## Results

### Participant characteristics

The analysis included 257 participants: 104 cases with a formal diagnosis of depression and 153 controls without a formal diagnosis. Most participants were female (203; 79.0%). High school completion was the highest reported educational level for 143 participants (55.6%), while 114 (44.4%) had at least a bachelor’s degree.

The overall mean PHQ-9 score was 8.47 (SD 7.62), and mean BMI was 23.78 kg/m^2^ (SD 4.54). There were 124 participants in the underweight/normal-weight group and 133 in the overweight/obese group. These groups included 44 and 60 cases, respectively. Depression status did not differ significantly between the two BMI groups (χ^2^=2.47; p=0.116). Participant characteristics are shown in Table 1.

**Table 1.** Characteristics of the study participants.

| **Characteristic** | **Total (n=257)** |
|---|---|
| Depression status, n (%) |  |
| Control: no formal diagnosis | 153 (59.5) |
| Case: formal diagnosis | 104 (40.5) |
| Sex, n (%) |  |
| Female | 203 (79.0) |
| Male | 54 (21.0) |
| Education, n (%) |  |
| High school graduate | 143 (55.6) |
| At least a bachelor’s degree | 114 (44.4) |
| BMI category, n (%) |  |
| Underweight | 18 (7.0) |
| Normal weight | 106 (41.2) |
| Overweight | 48 (18.7) |
| Obese | 85 (33.1) |
| BMI group, n (%) |  |
| Underweight/normal weight | 124 (48.2) |
| Overweight/obese | 133 (51.8) |
| Continuous variables, mean (SD) |  |
| PHQ-9 score | 8.47 (7.62) |
| Height, cm | 160.04 (7.83) |
| Weight, kg | 61.13 (13.92) |
| BMI, kg/m<sup>2</sup> | 23.78 (4.54) |

BMI, body mass index; PHQ-9, Patient Health Questionnaire-9; SD, standard deviation.

### Food-group intakes by depression status

Sweetened beverages were the only food group with a statistically significant overall difference between cases and controls. Mean intake was 2.14 units in cases and 1.53 units in controls, equivalent to approximately 214 and 153 g/day. The mean difference was 0.61 units, or approximately 61 g/day (95% CI 18–103; p=0.005).

Point estimates indicated higher mean intakes of fruits, milk and dairy products, and prepared foods, cakes, and snacks among cases, but the corresponding CIs included zero. The estimated difference in cereals and tubers was negative and also imprecise. None of the remaining food-group comparisons reached statistical significance (Table 2).

**Table 2.** Daily food-group intakes by depression status.

| **Food group** | **Controls (n=153), mean (SD)** | **Cases (n=104), mean (SD)** | **Mean difference, case − control (95% CI)** | **p-value** |
|---|---|---|---|---|
| Cereals and tubers | 5.36 (6.72) | 4.45 (4.26) | −0.90 (−2.25 to 0.45) | 0.188 |
| Legumes and nuts | 1.58 (1.88) | 1.68 (2.48) | 0.10 (−0.43 to 0.64) | 0.707 |
| Meat, poultry, and eggs | 2.84 (2.80) | 2.93 (3.21) | 0.10 (−0.65 to 0.84) | 0.800 |
| Fish and fish products | 0.77 (1.39) | 0.81 (1.72) | 0.05 (−0.34 to 0.43) | 0.812 |
| Vegetables | 3.25 (3.80) | 3.50 (5.08) | 0.26 (−0.83 to 1.35) | 0.643 |
| Fruits | 1.41 (1.79) | 1.97 (2.78) | 0.55 (−0.06 to 1.16) | 0.074 |
| Milk and dairy products | 1.15 (1.37) | 1.59 (2.89) | 0.45 (−0.16 to 1.05) | 0.144 |
| Oils and fats | 1.22 (1.35) | 1.33 (1.55) | 0.11 (−0.25 to 0.47) | 0.538 |
| Prepared foods, cakes, and snacks | 1.67 (1.62) | 2.09 (2.13) | 0.42 (−0.07 to 0.91) | 0.089 |
| Sweetened beverages | 1.53 (1.33) | 2.14 (1.88) | 0.61 (0.18 to 1.03) | 0.005 |

Intakes and mean differences are expressed in units of 100 g/day. Independent-samples t-tests were used, with Welch’s test when Levene’s p<0.05. CI, confidence interval; SD, standard deviation.

### Food-group intakes stratified by BMI group

The underweight/normal-weight group included 44 cases and 80 controls. Within this group, mean sweetened beverage intake was approximately 215 g/day in cases and 131 g/day in controls. The estimated mean difference was 85 g/day (95% CI 29–140; p=0.003).

The overweight/obese group included 60 cases and 73 controls. Mean sweetened beverage intake was approximately 213 g/day in cases and 178 g/day in controls, with an estimated difference of 35 g/day (95% CI −28 to 98; p=0.278).

No other food-group comparison reached statistical significance within either BMI group. These stratified comparisons suggested a larger sweetened beverage difference in the underweight/normal-weight group but did not independently establish that the associations differed between groups (Table 3).

**Table 3.** Daily food-group intakes by depression status, stratified by BMI group Panel A. Underweight and normal weight (n=124)

| **Food group** | **Controls (n=80), mean (SD)** | **Cases (n=44), mean (SD)** | **Mean difference, case − control (95% CI)** | **p-value** |
|---|---|---|---|---|
| Cereals and tubers | 5.40 (7.11) | 4.13 (3.71) | −1.27 (−3.55 to 1.01) | 0.272 |
| Legumes and nuts | 1.33 (1.36) | 1.57 (2.39) | 0.23 (−0.55 to 1.02) | 0.550 |
| Meat, poultry, and eggs | 2.88 (2.64) | 2.82 (2.29) | −0.06 (−1.00 to 0.88) | 0.901 |
| Fish and fish products | 0.54 (0.60) | 1.05 (2.37) | 0.51 (−0.22 to 1.24) | 0.170 |
| Vegetables | 3.18 (3.54) | 3.58 (5.23) | 0.40 (−1.36 to 2.17) | 0.649 |
| Fruits | 1.34 (1.46) | 1.92 (2.79) | 0.58 (−0.32 to 1.49) | 0.202 |
| Milk and dairy products | 1.33 (1.57) | 1.75 (2.62) | 0.42 (−0.45 to 1.28) | 0.337 |
| Oils and fats | 1.07 (1.42) | 1.49 (1.90) | 0.43 (−0.17 to 1.02) | 0.158 |
| Prepared foods, cakes, and snacks | 1.67 (1.50) | 2.11 (1.79) | 0.44 (−0.16 to 1.04) | 0.147 |
| Sweetened beverages | 1.31 (1.11) | 2.15 (1.65) | 0.85 (0.29 to 1.40) | 0.003 |
| Panel B. Overweight and obese (n=133) |  |  |  |  |
| **Food group** | **Controls (n=73), mean (SD)** | **Cases (n=60), mean (SD)** | **Mean difference, case − control (95% CI)** | **p-value** |
| Cereals and tubers | 5.31 (6.32) | 4.69 (4.63) | −0.62 (−2.56 to 1.32) | 0.527 |
| Legumes and nuts | 1.86 (2.30) | 1.77 (2.57) | −0.08 (−0.92 to 0.75) | 0.841 |
| Meat, poultry, and eggs | 2.79 (2.99) | 3.01 (3.75) | 0.23 (−0.93 to 1.38) | 0.701 |
| Fish and fish products | 1.01 (1.89) | 0.64 (1.00) | −0.37 (−0.91 to 0.16) | 0.170 |
| Vegetables | 3.32 (4.09) | 3.44 (5.01) | 0.13 (−1.44 to 1.69) | 0.873 |
| Fruits | 1.49 (2.10) | 2.00 (2.79) | 0.51 (−0.33 to 1.35) | 0.235 |
| Milk and dairy products | 0.95 (1.07) | 1.48 (3.08) | 0.53 (−0.30 to 1.37) | 0.206 |
| Oils and fats | 1.38 (1.26) | 1.21 (1.25) | −0.17 (−0.60 to 0.26) | 0.431 |
| Prepared foods, cakes, and snacks | 1.67 (1.76) | 2.08 (2.37) | 0.41 (−0.30 to 1.12) | 0.256 |
| Sweetened beverages | 1.78 (1.52) | 2.13 (2.05) | 0.35 (−0.28 to 0.98) | 0.278 |

Intakes and mean differences are expressed in units of 100 g/day. Independent-samples t-tests were used, with Welch’s test when Levene’s p<0.05. BMI, body mass index; CI, confidence interval; SD, standard deviation.

### Association between sweetened beverage intake and depression

In the unadjusted logistic model, each additional 100 g/day of sweetened beverages was associated with higher odds of depression (OR 1.27; 95% CI 1.08–1.49; p=0.004). The association remained after adjustment for BMI group, sex, and age (OR 1.29; 95% CI 1.09–1.52; p=0.003).

In the adjusted main-effects model, older age was associated with higher odds of depression (OR 1.49 per 10 years; 95% CI 1.13–1.97; p=0.005). The CIs for BMI group and sex included one.

The interaction model provided exploratory evidence that the sweetened beverage association differed by BMI group. The OR per additional 100 g/day was 1.63 (95% CI 1.21–2.18; p=0.001) in the underweight/normal-weight reference group. The interaction ratio of ORs was 0.70 (95% CI 0.49–0.99; p=0.046), indicating a smaller exposure OR in the overweight/obese group. Given the exploratory analyses and the interaction CI close to one, this finding was interpreted cautiously (Table 4).

**Table 4.** Logistic regression models for sweetened beverage intake and depression.

| **Model and term** | **OR (95% CI)** | **p-value** |
|---|---|---|
| Model 1: unadjusted |  |  |
| Sweetened beverages, per 100 g/day | 1.27 (1.08–1.49) | 0.004 |
| Model 2: adjusted main effects |  |  |
| Sweetened beverages, per 100 g/day | 1.29 (1.09–1.52) | 0.003 |
| BMI: overweight/obese vs. underweight/normal | 1.29 (0.75–2.23) | 0.357 |
| Sex: male vs. female | 0.53 (0.27–1.04) | 0.065 |
| Age, per 10 years | 1.49 (1.13–1.97) | 0.005 |
| Model 3: adjusted with interaction |  |  |
| Sweetened beverages, per 100 g/day, in underweight/normal weight | 1.63 (1.21–2.18) | 0.001 |
| BMI: overweight/obese vs. underweight/normal, at zero beverage intake | 2.48 (1.07–5.78) | 0.035 |
| Sweetened beverages × BMI group | 0.70 (0.49–0.99) | 0.046 |
| Sex: male vs. female | 0.51 (0.26–1.01) | 0.052 |
| Age, per 10 years | 1.49 (1.13–1.97) | 0.005 |

All models included 257 participants, comprising 104 cases and 153 controls. The outcome was formal diagnosis of depression. P-values are Wald tests. BMI, body mass index; CI, confidence interval; OR, odds ratio.

## Discussion

### Principal findings

In this exploratory case–control study, participants with a formal diagnosis of depression reported higher sweetened beverage intake than controls without a formal diagnosis. The association remained after adjustment for BMI group, sex, and age. None of the other nine food groups showed a statistically significant overall difference between cases and controls. Exploratory analyses suggested that the sweetened beverage association was stronger among underweight or normal-weight participants than among those who were overweight or obese.

These findings identify a dietary difference within the study sample rather than establish a causal effect. The adjusted association was independent of the variables included in the model in a statistical sense, but it was not necessarily independent of total dietary intake, socioeconomic circumstances, treatment, or other unmeasured factors.

### Sweetened beverage intake in the context of diet and depression

The findings add a food-group-specific observation to the broader literature linking dietary patterns with depressive outcomes. Previous observational evidence has focused substantially on overall diet-quality indices, which capture combinations of foods and behaviors rather than a single beverage category. The present result should therefore not be interpreted as a direct test of those dietary patterns. Consistent with the direction of the present finding, a meta-analysis of observational studies reported a modestly higher risk of depression with greater sugar-sweetened beverage consumption,^7^ and prospective cohort studies have examined sugar and sweetened-beverage intake in relation to later depression.^8–10^ A systematic review and meta-analysis also reported that higher total sugar consumption was associated with a higher risk of depression.^11^ Findings appear to differ by beverage type, however: in a Japanese cohort, sweetened coffee was associated with higher risk whereas black coffee was associated with lower risk,^10^ and in a US cohort of older adults the association was most evident for artificially sweetened beverages,^9^ distinctions that a weight-based beverage measure cannot capture.

Several proposed pathways could connect dietary behavior with mental health, including metabolic regulation, inflammation, oxidative stress, and gut–brain signaling. However, the study did not measure these mediators and cannot determine whether any such pathway explains the observed association.

The exposure was the estimated weight of sweetened beverages consumed, not measured sugar intake. Beverages grouped under this label may differ in sugar concentration, energy content, caffeine, and other ingredients. Consequently, the result cannot be attributed specifically to sugar, caffeine, or any individual beverage component without further item-level analysis.

Reverse causation is also plausible. Depression, associated changes in appetite or daily routines, and treatment may influence beverage preferences and consumption.^12^ The observed association could therefore reflect dietary behavior following illness, a preceding exposure, shared determinants, or some combination of these processes.

### BMI and the exploratory interaction

The sweetened beverage difference was larger in the underweight/normal-weight group. The regression interaction was consistent with a smaller exposure OR among overweight/obese participants, but the evidence was limited: the interaction p-value was 0.046, and its CI approached the null value. This result arose alongside multiple food-group and subgroup comparisons and should be treated as hypothesis-generating.

A statistically significant association within one stratum and a non-significant association within another does not, by itself, demonstrate effect modification. The interaction term provides the relevant model-based comparison, but it does not remove concerns about multiplicity, model specification, or residual confounding. Nor does the finding demonstrate that overweight or obesity protects against any potential consequence of sweetened beverage consumption.

BMI was measured in relation to current participant characteristics rather than established as preceding both diet and depression. Its role may therefore be complex.^13^ Depending on the temporal relationships, BMI could represent a confounder, an intermediate variable, or a consequence of illness or treatment. Adjustment and stratification by BMI should not automatically be interpreted as isolating a causal dietary effect.

### Interpretation of the other food groups

The remaining food-group comparisons did not provide clear evidence of differences between cases and controls. However, non-significant results should not be interpreted as proof of equivalent intake or absence of an association. Several estimates were imprecise, and the broad food groups may combine products with different nutritional characteristics.

For example, the oils and fats category does not quantify total dietary fat, while prepared foods, cakes, and snacks do not constitute a validated ultra-processed food measure solely by virtue of their group label. Similarly, milk and dairy products or legumes and nuts do not distinguish fermented from non-fermented items. The present analysis therefore does not establish nutrient-specific, fermentation-specific, or processing-specific associations.

The mean fruit and vegetable intakes also address a different question from the proportion of participants meeting a combined daily intake threshold. Continuous group means cannot be used to infer how many individuals met a recommendation without the corresponding participant-level classification.

### Clinical characterization and generalizability

Formal diagnosis and recent depressive symptoms describe related but distinct aspects of clinical status. The overall PHQ-9 mean summarizes symptom burden across the combined sample but does not establish the severity distribution within either diagnostic group. It also cannot determine whether cases were experiencing an active episode, partial improvement, or remission.

Controls were defined by the absence of a formal diagnosis in the analysis. This definition should not be interpreted as proof that all controls were free of depressive symptoms or undiagnosed depressive disorders. Diagnostic ascertainment and the eligibility rules actually applied are therefore important to the interpretation of the comparison.

The sample was predominantly female and drew cases and controls from different recruitment settings. Its characteristics may not represent those of the wider Tangerang population. The observed case proportion is a feature of the analytic sample, not an estimate of depression prevalence.

### Strengths and limitations

A strength of the study is its assessment of multiple locally relevant food groups within one participant sample. Presenting the complete set of food-group comparisons provides a broader dietary description than focusing only on the statistically significant finding. The analysis also examined the sweetened beverage association using both descriptive comparisons and regression models.

Several limitations warrant consideration. First, the case–control design and assessment of recent diet do not establish whether intake preceded diagnosis. Reverse causation and treatment-related changes in eating behavior remain possible.

Second, recruitment of cases from psychiatric outpatient services and controls through university and community networks may introduce selection and comparability concerns. Differences associated with recruitment setting cannot necessarily be removed by adjustment for age, sex, and BMI.

Third, FFQ-based estimates are subject to recall and portion-size errors.^5^ Some food-group standard deviations were large relative to their means, indicating substantial variability and motivating careful assessment of distributions and influential observations. The reported mean comparisons do not fully describe individual intake distributions.

Fourth, the adjusted models included a limited set of covariates. Total energy intake, other dietary exposures, socioeconomic circumstances, physical activity, smoking, alcohol use, and depression treatment were not included in the presented models. Residual confounding therefore remains possible.

Fifth, the regression models expressed sweetened beverage intake as a linear term per 100 g/day on the log-odds scale. The presented results do not establish a threshold or exclude a nonlinear relationship.

Finally, multiple food-group comparisons and subsequent subgroup and interaction analyses increase the possibility of chance findings. The BMI interaction should be interpreted particularly cautiously, and the analyses should not be presented as confirmatory evidence for a prespecified subgroup effect.

## CONCLUSIONS

Higher sweetened beverage intake was associated with a formal diagnosis of depression in this Tangerang case– control sample, including after adjustment for BMI group, sex, and age. Other food-group comparisons were inconclusive. Exploratory evidence suggested a stronger sweetened beverage association among underweight or normal-weight participants, but this interaction requires confirmation. The findings do not establish causality or support BMI-specific dietary recommendations. Longitudinal research with more detailed dietary, clinical, and confounder assessment is needed to clarify temporal relationships.

## Data Availability

All data produced in the present study are available upon reasonable request to the authors

## ACKNOWLEDGEMENT

The authors thank all study participants, and the staff of the psychiatric outpatient services and the university and community partners who helped with recruitment and data.

## AUTHOR CONTRIBUTIONS

DC: conceptor, investigation, methodology, validation, writing original draft, writing review; AS: conceptor, methodology, validation, writing original draft, writing review; NS: investigation, writing original draft, writing review; RS: data curation, formal analysis, investigation, validation, writing original draft, writing review; DR: conceptor, investigation, methodology, supervision, validation, writing original draft, writing review. All authors reviewed and approved the final manuscript.

## DATA AVAILABILITY

The datasets analysed during the current study are available from the corresponding author on reasonable request, subject to participant consent, ethics approval, and institutional data-governance requirements.

## PRIOR DISSEMINATION

This work has not been published previously and is not under consideration elsewhere.

## USE OF AI-ASSISTED TOOLS

Generative artificial intelligence (Claude, Anthropic) was used to assist with translating and language-editing this manuscript under the authors’ supervision throughout. All data interpretation, analysis, and final scientific and editorial decisions were made exclusively by the authors, and the underlying research data were generated and analyzed by the authors without AI involvement.

